# Temporal and Thematic Analysis of Promotional Waterpipe-Related Posts on Twitter/X in the US

**DOI:** 10.1101/2024.10.17.24315663

**Authors:** Puhua Ye, Mengwei Wu, Yiwei Han, Yuka Shimazaki, Jennifer Cornacchione Ross, Erin L. Sutfin, Dongmei Li, Zidian Xie

## Abstract

**Introduction:** Waterpipe tobacco smoking (WTS), also known as hookah, shisha, or narghile, is particularly popular among young people in the United States (US). WTS poses serious health risks similar to those of cigarette smoking.

**Methods:** Using the Twitter/X streaming API (Application Programming Interface), we collected 4,853,562 tweets between March 9, 2021, and March 14, 2023, using waterpipe-related keywords, such as “hookah” and “waterpipe”. After geographical filtering to identify tweets from the US and keyword filtering for the promotional content, we identified 23,803 promotional waterpipe-related tweets. We examined trends in the posting time of these promotional waterpipe-related tweets and identified prevalent topics from these tweets using the BERTopic (Bidirectional Encoder Representations from Transformers) modeling.

**Results:** The number of promotional waterpipe-related tweets showed an overall decreasing trend during the study period. The posting of promotional waterpipe-related tweets was more active later in the day. Major topics in the promotional tweets included “Promotion from hookah lounges and online hookah business" (63.97%, 15,227/23,803), "promoting hookah parties and events" (32.26%, 7,679/23,803), and "promoting engineered and durable hookah products" (3.77%, 897/23,803). Twitter/X accounts posting waterpipe-related promotional content have substantial variations in the number of relevant tweets (mean = 2.28, SD = 12.22) and followers (mean = 5,937, SD = 76,770).

**Conclusions:** This study demonstrates a significant social media activity in promoting waterpipe tobacco smoking. Our findings underscore the urgent need to regulate the promotional content of WTS on social media and promote public health education messages on social media to counteract the promotion of WTS.

## Introduction

Waterpipe tobacco smoking (WTS), also known as hookah, shisha, or narghile is a traditional smoking device that originated in India over a millennium ago and has recently gained popularity in the United States (US) (1). In 2021, approximately 0.9% of US adults, which equates to an estimated 2.2 million people, reported currently using hookah (2). Among those who use waterpipe, young people, especially college students, make up the majority of users (3). Around 8-17% of college students currently use waterpipe tobacco, and about 35-64% have tried it at least once. A recent systematic review showed that waterpipe use among young people in the US is increasing by up to 1% annually (4). Among university students in the US, waterpipe tobacco smoking is the second most prevalent form of tobacco use, following cigarettes (5). Another study suggests that college students are at a higher risk of smoking waterpipe and there is also a notable knowledge gap and misperception regarding the potential harm of hookah, particularly among young, college-educated, and never-smokers (6).

Most waterpipe users believe that smoking waterpipe tobacco is less harmful than other forms of tobacco use (6). This misconception is particularly prevalent among college students (6–8). In a study of college students, less than 37% of them correctly answered any questions about the toxins produced by a 45-minute waterpipe session compared to a five-minute cigarette session (8). Some studies report that college students mistakenly believe that waterpipe smoking is less harmful than cigarette smoking due to the perceived filtering effect of water, the social and intermittent use patterns, and the smooth, aromatic smoke (9, 10). However, in reality, one session of waterpipe tobacco smoking consistently exposes users to larger volumes of smoke and higher levels of toxic substances, such as nicotine, tar, and carbon monoxide (CO) compared to smoking a single cigarette (11). WTS is also likely associated with lung cancers, as well as respiratory diseases, low birth weight, metabolic syndrome, cardiovascular disease, and mental health issues (1, 12, 13). Additionally, the manner in which waterpipe tobacco is used, along with the common practice of sharing the mouthpiece, leads to additional health risks compared to other tobacco products. Sharing the mouthpiece during WTS can transmit pathogens such as viruses, bacteria, and fungi, increasing the risk of diseases like hepatitis C, tuberculosis, peptic ulcers, pneumonia, and periodontal diseases (1, 14–17).

An important factor contributing to the misconceptions about WTS and its widespread popularity is the influence of media (18). In the early 1990s, satellite TV broadcasts from cafés and restaurants during Ramadan helped to advertise and popularize the waterpipe, making it more familiar to people in the Middle East (18). Today, social media has become a major avenue for promoting waterpipe (19, 20). There is substantial evidence that exposure to tobacco marketing and promotions, including on social media, is associated with tobacco product initiation and use, reduced harm perceptions, and increased positive tobacco product attitudes (21, 22). Promotions are used to increase interest in and sales of products. However, research has also demonstrated social media to be a promising channel for disseminating risk messages about WTS by counteracting the impact of promotional content (23). Therefore, understanding how waterpipe is promoted on social media is crucial to inform policies about marketing restrictions and identifying targets for risk messaging.

Twitter/X is a widely-used social media platform with over 500 million monthly active users worldwide. In the US, it has more than 50.5 million monthly active users in 2023. Around 60% of Twitter/X users are youth and young adults aged 13 – 34 (24). Twitter/X data have been utilized to examine public perceptions and discussions regarding tobacco products, including waterpipe (25–28) and related regulatory policies (29–31). This study aims to provide important insights into how waterpipe is promoted on social media. By analyzing promotional waterpipe- related tweets, this research seeks to identify the main themes and trends in waterpipe promotion on Twitter/X. The findings could inform future regulatory measures, such as potential promotional marketing restrictions, and public health policies to reduce waterpipe use and associated health risks.

## Materials and Methods

### Ethics Statement

This study was reviewed and approved by the Research Subjects Review Board of the Office for Human Subject Protection at the University of Rochester (STUDY00006570). Patient consent for publication is not required as the data were analyzed anonymously.

### Data Collection and Preprocessing

Twitter/X data related to waterpipe were collected in real-time using the Twitter/X streaming API (Application Programming Interface) between March 9, 2021, and March 14, 2023, using waterpipe-related keywords such as “hookah” and “waterpipe” (26, 32). The initial dataset contains 4,853,562 tweets. To focus on tweets from the US we applied a geographical filter based on the user location metadata. Specifically, we used the names and abbreviations of the 50 US states and major US cities to identify tweets originating from the US (33, 34). Following geographical filtering, we further filtered the dataset to identify tweets related to waterpipe smoking. A comprehensive list of keywords was used, including terms such as "waterpipe," "hookah," "shisha," "narghile," "argileh," "goza," and "hubble-bubble." These keywords were identified within the tweet text. Furthermore, several data-cleaning steps were performed. For example, duplicate tweets and retweets have been identified and removed by checking for tweets starting with "RT @" and comparing the text of tweets to ensure uniqueness.

Tweets were classified into promotional and non-promotional categories using a rule-based approach. We defined ’promotional’ tweets as those that explicitly or implicitly promote waterpipe use, including tweets from waterpipe brands, online retailers, hookah bars/lounges, and consumers engaging in promotional activity. This classification was based on the presence of specific keywords indicative of promotional content in the tweet text, user screen name, and username. Keywords included terms such as "dealer," "deal," "promotion," "sale," "discount," "store," "promo," and others (27). The final dataset consisted of 323,347 waterpipe-related tweets from the US, with 299,544 non-promotional tweets and 23,803 promotional tweets.

### Data Analysis

We conducted a temporal analysis of the tweets to understand the distribution of promotional waterpipe-related tweets over time. The timestamp (adjusted for the local time zone) of each tweet is in the ‘timestamp_ms’ field. We grouped the tweets based on this timestamp to analyze their frequency by hour, day, and month. This clustering allowed us to identify periods of high and low activity. We visualized the results using Matplotlib and Tableau to create time series plots and heatmaps.

For user analysis, we collected the unique user ID, follower count, and the number of tweets posted by each Twitter/X user. The follower count for each user was determined by extracting the highest follower count reported in the relevant tweets by this user. The total number of tweets posted by each user was calculated by aggregating the tweets during our study period based on the user ID.

### Topic Modeling Analysis

We utilized BERTopic (Bidirectional Encoder Representations from Transformers) to identify popular topics in promotional waterpipe-related tweets. BERTopic is a topic modeling technique that generates the embedding (numerical representations of the text data in a high-dimensional space that capture the semantic meaning of the words and phrases) with a pre-trained language model, clusters these embedding, and generates topic representations with the class-based TF- IDF procedure (35). To ensure consistency in the preprocessing steps, we converted all characters to lowercase and lemmatized all words using WordNetLemmatizer. Additionally, we removed stop words (e.g. a, an, and, but, in), punctuation, web links, and emojis using the Natural Language Toolkit (NLTK) and Regular Expression (RE). We initially applied K-Means clustering to group the tweets and set the ‘hdbscan_model’ parameter to the model trained with K-Means. We experimented with the number of clusters from three to ten and determined the optimal number of clusters using coherence scores and visualization maps to observe the intertopic distance.

## Results

### Temporal Analysis of Promotional Waterpipe-Related Tweets

We collected a total of 4,853,562 tweets from March 9, 2021, to March 14, 2023 using waterpipe-related keywords, of which 323,347 waterpipe-related tweets were identified as originating from the United States. There were 299,544 non-promotional waterpipe-related tweets (92.64%) and 23,803 promotional waterpipe-related tweets (7.36%). In this study, we focused on promotional waterpipe-related tweets. Fig 1 illustrates the monthly frequency of promotional waterpipe-related tweets, revealing distinct trends over the study period. From April 2021 to July 2021, there was a general upward trend, followed by an overall gradual decrease.

**Figure 1.**
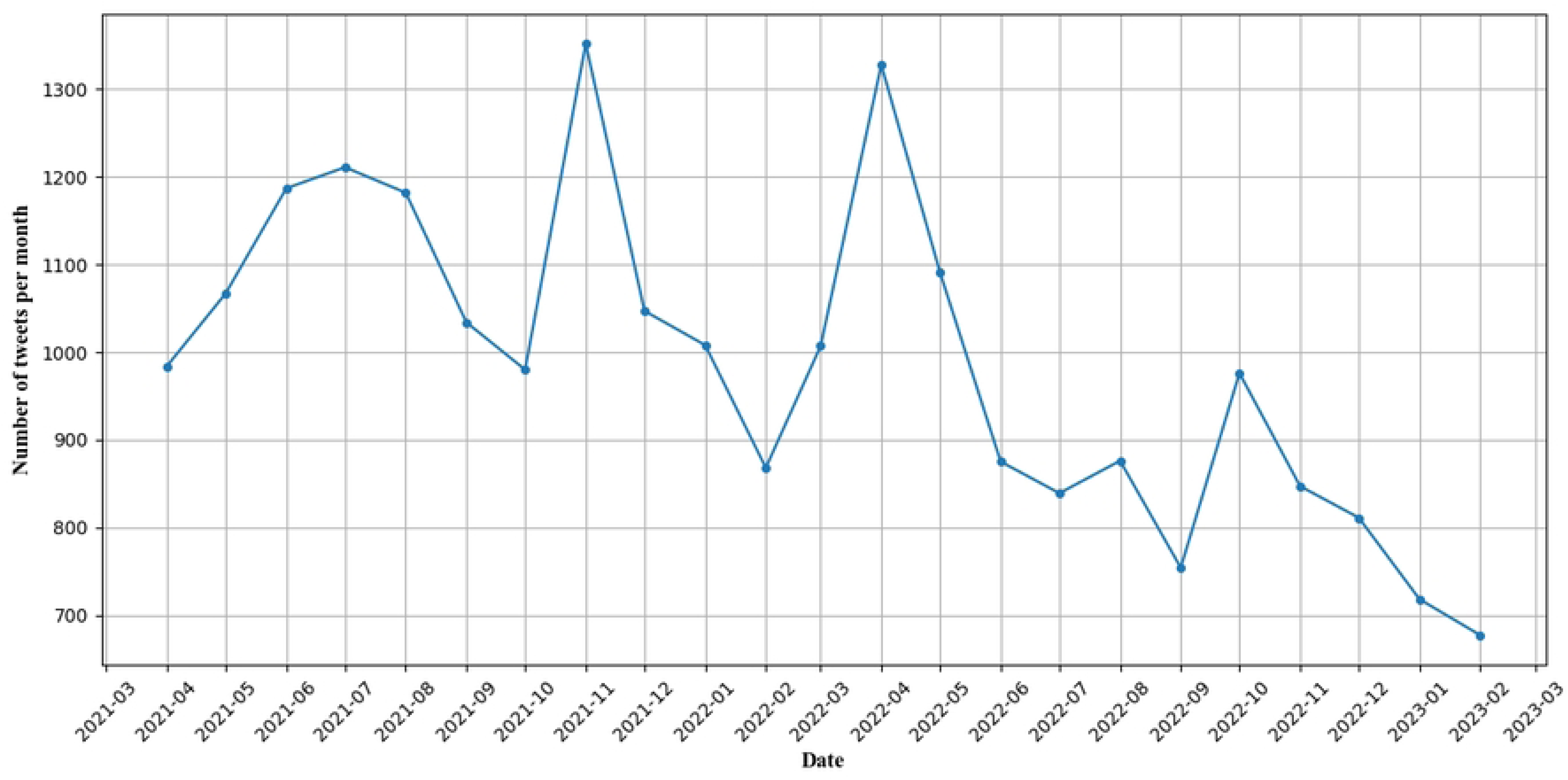
**Temporal Trend in Promotional Waterpipe-Related Tweets.**

However, we observed two noticeable peaks, one is in November 2021 and the other one is in April 2022. From S1 Figure, we can see that the overall trend in the number of users is similar to the trend in the number of tweets except that in November 2021 the number of users did not show a noticeable peak.

Fig 2 presents the distribution of promotional waterpipe-related tweets by hour in the day and day of the week, with a color gradient representing tweet volume—darker shades indicate higher activity while lighter shades represent lower activity. The overall activity patterns show some variation in tweet frequency throughout the day and the week. The tweet volume is generally lower during the early morning and increases as the day progresses, peaking in the late afternoon and evening. The highest activity is observed on Tuesdays at 6:00 PM, with 327 tweets. Across most weekdays, particularly from Tuesday to Thursday, the high posting activity consistently appears from 4:00 PM to 8:00 PM. Those weekdays, especially Tuesday to Thursday, experience higher posting activity than the weekend.

**Figure 2.**
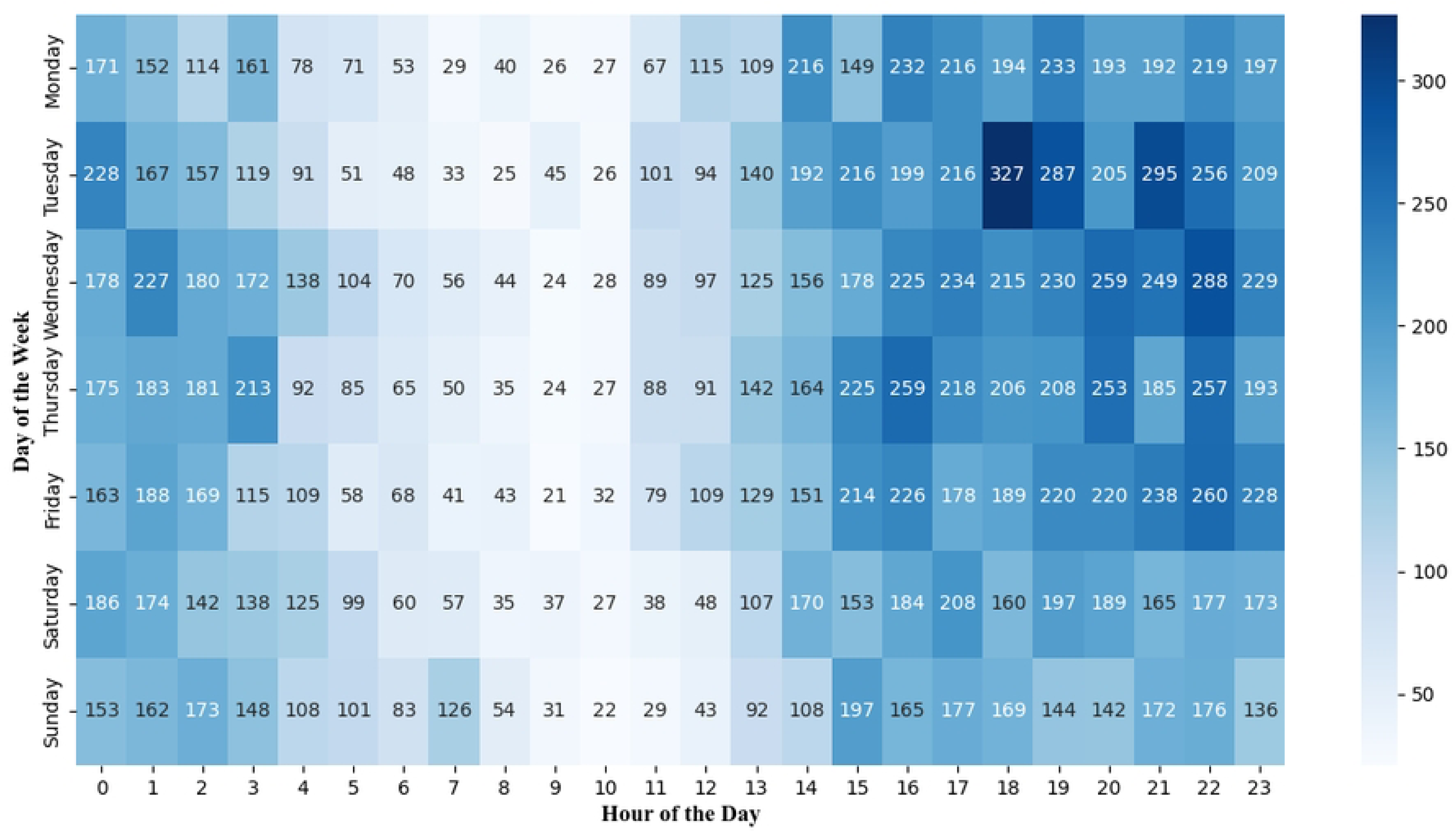
**Number of Promotional Waterpipe-Related Tweets Posted at Different Times in a Day and Days of the Week.**

### Analysis of Twitter/X Accounts for Promoting Waterpipe

From 23,803 promotional waterpipe-related tweets, we have identified 10,524 unique Twitter/X accounts. The median number of promotional waterpipe-related tweets per Twitter/X account is 1, with a range from 1 to 690. In our analysis of 10,524 accounts with available follower counts, the average number of followers per account was 5,937 (the median = 768), with a range from 0 to 4,008,382. The follower counts varied significantly (SD=76,770), with 3.2% of accounts having fewer than ten followers and 5.7% having more than 10,000 followers.

S1 Table provides the top ten Twitter/X users with the most promotional waterpipe-related tweets during our study period. Among these top ten Twitter/X users, five accounts belong to hookah business owners who use Twitter/X to advertise their online waterpipe shops, four accounts are associated with hookah lounges promoting events at their establishments, and one account falls into the "other" category, focusing on teaching people how to smoke hookah. The number of tweets posted by these users varied significantly, ranging from 204 to 690 tweets. In addition, we observe a wide range of follower counts among the users, from as few as three followers to as many as 5,884 followers.

### Major Topics Discussed in Promotional Waterpipe Related Tweets

To understand the main themes in the promotional waterpipe-related tweets, we performed a topic modeling analysis using BERTopic. As shown in Table 1, three major topics were identified, including “Promotion from hookah lounges and online hookah business” (63.97%, 15,227/23,803), “Promoting hookah parties and events that offer hookah” (32.26%, 7,679/23,803), and “Promoting engineered and durable hookah products” (3.77%, 897/23,803).

**Table 1.** Major Topics Discussed in Promotional Waterpipe-Related Tweets.

| Major topic (percentage, n/N) | Description | Keywords | Example tweets |
| --- | --- | --- | --- |
| Topic 1: Promotion from hookah lounges and online hookah business. (63.97%, 15,227/23,803) | Tweets promote various hookah lounges and their offerings, emphasizing the events such as special nights, often featuring different shisha flavors and special deals, and various hookah business and their products, emphasizing the quality and discounts. | hookah, smoke, lounge, come, store, night, flavor, free, like, shisha | 1. “BOOM: New 21+ Friday Night Wave For The People \$4 Sex On The Beach \$7 Henny & Patron \$20 Hookah Catered Food LITERALLY NO COVER CHARGE MEANING FREE ENTRY ALL NIGHT NO GAMES NO GIMMICKS #WeBackOutsideFridays Culture Lounge”<br>2. “New Product - Delta 8 Shisha \$39.99 Available Now in 2 Flavors - Mighty Mint and Pink Lemonade” |
| Topic 2:<br>~~Promoting parties~~<br>and events that offer hookah. (32.26%, 7,679/23,803) | ~~Tweets advertise parties and gatherings, often~~<br>offering free entry, complimentary food, and shisha. These events are typically scheduled for weekends, creating a lively and social atmosphere for attendees. | free,<br>hookah,<br>night,<br>shisha,<br>party, come,<br>tonight,<br>drink,<br>happy,<br>Saturday | 1. “PLS RT ☺ Brunch + ~~Mimosas + Drink Specials~~ + Hookah + #SunDAYFUNday #BrunchaholicsAnonymous Reservations/Info:”<br>2. “FTS ITS HOMECOMING DAY PARTY March 13, 2021 2pm -7pm Greeks \$5 all day Get the pre-sales now Hookah and Live Dj ——— Txt Number on flyer for the addy #ASUTwitter #FVSU” |
| Topic 3:<br>~~Promoting~~<br>engineered and durable hookah products. (3.77%, 897/23,803) | ~~The tweets emphasize the advanced design and~~<br>quality of the waterpipe tobacco, making it a standout product in the market. | revolutionar<br>y,<br>engineered,<br>durable,<br>smooth,<br>waterpipe,<br>available,<br>easily,<br>whatsa,<br>calhounmem<br>orial, suite | 1. “Engineered to be the ~~smoothest, most durable,~~ and revolutionary waterpipe available!”<br>2. “Our hookah’s are perfect for those at home date nights, Valentine’s Day, girls nights & baecations! All hookah’s come with shisha, coals & hookah tips! We also sell custom shisha mixes and jolly rancher candy hookah tips!” |

Figure 3 shows the distribution of these main topics over the study period. We observed that the topic of promoting hookah lounges and shisha flavors had the highest number of tweets throughout the study period, peaking several times. The topic of promoting hookah parties and events showed periodic peaks, particularly during months with major holidays. The topic of promoting engineered and durable hookah products had the lowest volume but maintained a steady presence over time. From S2 Figure, we can observe that the proportion of Topic 1 consistently remains between 60% and 70%, maintaining a dominant presence during the study period. Noticeable, Topic 2 shows several significant increases, such as in November 2021, April 2022, and October 2022. These peaks also coincide with the overall tweet trend peaks during those periods.

**Figure 3.**
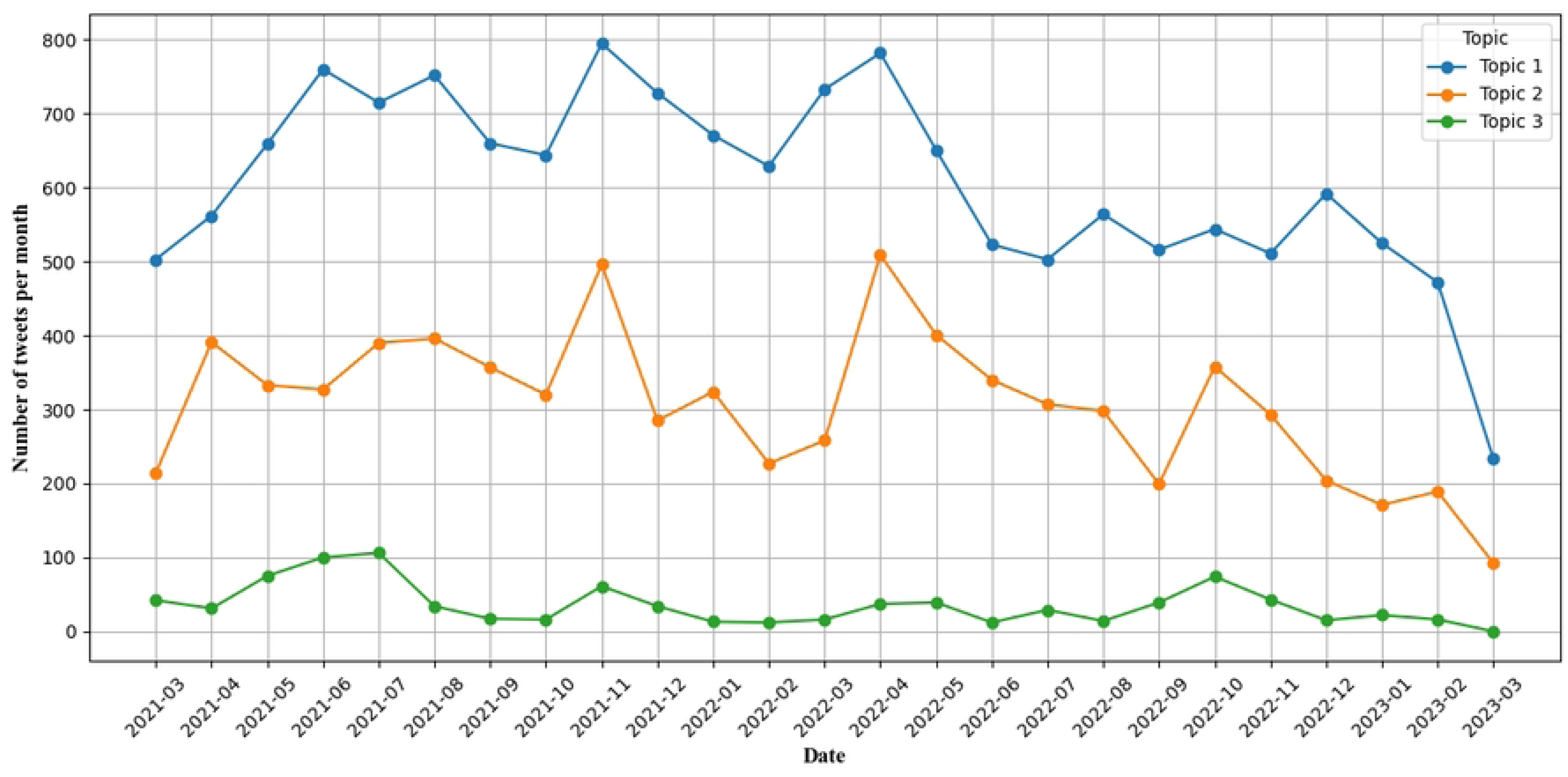
Major Topics in Promotional Waterpipe-Related Tweets over Time. Topic 1: Promotion from hookah lounges and online hookah business; Topic 2: Promoting parties and events that offer hookah; Topic 3: Promoting engineered and durable hookah products.

## Discussion

Using Twitter/X data from March 9, 2021, to March 14, 2023, this study examined waterpipe tobacco-related promotions on social media. The temporal trend analysis indicates an initial increase in the number of promotional tweets until July 2021, followed by a general decline except for two significant peaks in November 2021 and April 2022. The topic modeling analysis identified three primary themes in the promotional tweets, including hookah lounges promoting their establishments through discounts and events, party events or other venues emphasizing the availability of hookah as part of their offerings, and the promotion of specific hookah products. The frequency of promotional tweets did not appear to be related to the number of followers, suggesting that businesses of various sizes, irrespective of their social media following, utilize Twitter/X to promote waterpipe smoking.

During the study period, two significant peaks in the number of promotional waterpipe-related tweets appeared in November 2021 and April 2022. After manually reviewing the tweet content from these two months, we found that The November 2021 peak corresponded with heightened promotional activity, while the April 2022 peak coincided with a rising trend of enjoying the waterpipe tobacco during brunch, exemplified by tweets such as: *"#TheBrunchMarket returns to #BeatsBrunchandBubbly TODAY from 12pm to 4pm at RainDogs in #FivePointsJax!• Beats by EnerJi• Brunch by: High Elevated Eatz• Bubbly: $12 AYCD #BottomlessMimosas• Hookah Service• Featuring Local Vendors•: Come thru &amp; catch the vibes!!"* and *"Brunch n Boozy Sundays 12pm - 6pm, Section 6 People with hookah &amp; bottomless mimosas $175, High Top 4 People with hookah &amp; bottomless mimosas $125, Section 6 People 1 Bottle $250"*. The two peaks are consistent with the similar peaks in Topic 1 (promotion of hookah lounge) and Topic 2 (promotion of parties with hookah). There might be two main factors contributing to the peak in November 2021. First, we observed that some hookah lounges began engaging in direct marketing on social media by responding to users’ tweets to promote their services. In November, the number of promotional tweets that began with "@", which means direct replies to specific users, increased from 112 in the previous month to 465 tweets. This finding aligns with one previous study showing that direct marketing is a crucial channel for tobacco companies (36). For instance, 16 million (7.1%) US adults reported receiving e-cigarette promotions via mail or email between 2013 and 2014 (37). In addition, the presence of Black Friday and Thanksgiving holidays in November led to an increase in tweets from hookah lounges offering discounts and event organizers promoting free hookah at parties. Another study on e-cigarette marketing on Twitter/X reported that about a third of all commercial e-cigarette tweets during two months (May–June 2012) contained mentions of prices or discounts (38). We suspect that another surge in promotional tweets in April 2022 might partially result from the celebration of the Spring season. Specifically, there was a notable rise in tweets promoting enjoying waterpipe tobacco during brunch.

We noticed a surge of promotional waterpipe tobacco-related tweets in the late afternoon and evening, especially Thursday and Tuesday. Several factors may explain the observed patterns. The increased activity during late afternoons and evenings on weekdays could be attributed to users being more active on social media after working or school hours, making these times optimal for promoting WTS. The high volume of tweets on Thursdays might be due to hookah bars or lounges posting announcements for upcoming weekend events to attract customers.

Similarly, the peak on Tuesdays could be related to promotional strategies targeting early week engagement to boost midweek and weekend attendance at venues. To counteract the impact of waterpipe tobacco promotional activity on social media, the waterpipe tobacco prevention messages should be encouraged during these peak days and times of promotional tweets, ensuring they reach the target audience when promotional activity is at its highest.

The topic analysis revealed that waterpipe-related promotions predominantly occurred through hookah lounges and other social event organizers, which is consistent with Ziyad’s study showing that promotional posts often highlighted events with 37.4% promoting activities held at hookah lounges or bars and 22.2% featuring general events where hookah was also available (39). In contrast, traditional tobacco products often focus on the product’s attributes, such as quality, flavor, and health claims (e.g., e-cigarettes marketed as safer alternatives to smoking) (40). The difference in the promotion strategy between waterpipe and other tobacco products can be attributed to the unique usage method of waterpipes and its inherent social attributes. Unlike portable tobacco products, waterpipes consist of a head (bowl), a vertical stem (body), and a flexible hose with a mouthpiece. The waterpipe tobacco is heated by burning charcoal, producing smoke that the user inhales through the hose (41), a setup that is less convenient for on-the-go use compared to cigarettes or e-cigarettes. In addition, WTS has a strong social component (7, 42–44). A previous study involving café patrons and internet forum users found that one of the most common perceived positive attributes of waterpipe tobacco smoking was the opportunity to socialize with friends, with 79.6% of participants highlighting this aspect (45). This social nature makes waterpipe lounges and events ideal venues for promoting WTS, differentiating it from other tobacco products often marketed based on individual usage and convenience. The direct and active promotion of WTS on social media by hookah lounges and other businesses can potential promote the waterpipe tobacco use, which should be closely monitored and regulated by public health authorities to protect public health.

Among those Twitter/X users posting promotional waterpipe-related tweets, including the top 10 user accounts, there was a significant variation in the number of waterpipe-related tweets and followers. This finding aligns with a previous study on e-cigarettes on Instagram, which revealed that the number of followers varied greatly for accounts promoting e-cigarettes (38). These results indicate that both large and small businesses are actively utilizing social media platforms to promote tobacco products, including WTS. This trend highlights the diverse range of social media users engaged in marketing activities, regardless of their follower base size. Moreover, in the top 10 user accounts, there is an equal representation of hookah lounges and hookah online retailers, illustrating that waterpipe tobacco promotion involves both online sales, similar to other tobacco products, and a substantial presence from hookah lounges. The widespread use of social media for promoting waterpipe tobacco smoking underscores the need for government regulation to effectively monitor and manage these activities.

Our study has several limitations. First, using the keywords to filter and classify promotional vs. non-promotional tweets may have inherent biases and inaccuracies. Future research could use machine learning or deep learning methods to improve the accuracy of tweet classification.

Second, our data collection was limited to Twitter/X, so our findings may not reflect promotional activities on other social media platforms due to different user behaviors and demographics.

Future work is warranted to investigate the waterpipe promotional activity on other social media platforms (such as Instagram and TikTok) that are more likely to target at the youth and young adults. Third, although we filtered for tweets from the United States, some tweets might have been inaccurately geo-tagged or lacked geolocation information, leading to potential misrepresentation of the geographic distribution of promotional activities. Fourth, when analyzing the user types of Twitter/X users promoting waterpipe, we only categorized the top 10 users with the highest follower counts, which represents a biased sample and may not reflect the full spectrum of users promoting waterpipe tobaccos. Future studies should aim to analyze all users promoting waterpipe tobaccos to better understand these Twitter/X users. Last, Twitter/X does not provide demographic information (such as age, gender, and race/ethnicity) on Twitter/X, making it impossible to determine the demographics of followers of waterpipe tobacco-promoting accounts. Future studies should consider addressing these limitations by employing more advanced classification techniques and expanding the scope to include multiple social media platforms. In addition, with the rapid change in the waterpipe tobacco marketing, continuous monitoring of waterpipe tobacco promotion on social media become necessary.

## Conclusion

This study examined the promotion activity of waterpipe tobacco smoking (WTS) on Twitter/X. The analysis indicated an initial increase in promotional tweets until July 2021, followed by a general decline. The tweets mainly focus on hookah lounges promoting their establishments through discounts and events, party events, or other venues emphasizing the availability of hookah, and the promotion of specific hookah products. User analysis revealed that businesses of various sizes, irrespective of the size of their social media followers, utilize Twitter/X to promote waterpipe tobacco smoking. These findings highlight the strategic use of social media platforms like Twitter/X to promote waterpipe tobacco smoking, particularly among younger audiences, and emphasize the necessity for targeted public health interventions to address this issue.

Continuous monitoring of promotional activities and understanding the factors driving these trends can inform policy decision-making and public health initiatives, such as regulating the content of online advertisements of waterpipe tobacco smoking. By addressing these promotional strategies, policymakers can better regulate waterpipe tobacco marketing and reduce its associated health risks.

## Data Availability

The data underlying the results presented in the study are available from Twitter/X (https://x.com).

## Acknowledgements

None.

## Contributions

ZX, DL: conceived and designed the study. PY, MW, YH, YS, ZX, DL: analyzed the data. PY, JCR, ELS, ZX, and DL: assisted with interpretation of analyses and wrote and edited the manuscript.

## Disclaimer

The content is solely the responsibility of the authors and does not necessarily represent the official views of the NIH or the FDA.

## Data Availability

The data present in this study can be requested from the corresponding author.

## Ethics approval

This study has been reviewed and approved by the Office for Human Subject Protection Research Subjects Review Board (RSRB) at the University of Rochester (Study ID: STUDY00006570).

## Supporting Information

**S1 Fig. Temporal trend in promotional waterpipe-related users**

**S2 Fig. Proportion of major topics in promotional waterpipe related tweets over time.** Topic 1: Promotion from hookah lounges and online hookah business; Topic 2: Promoting hookah parties and events; Topic 3: Promoting engineered and durable hookah products.

S1 Table. Top Twitter/X accounts with the most promotional waterpipe-related tweets.

